# Comparison of selected nutritional status and disease biomarkers in omnivores, flexitarians, pescatarians, vegetarians, and vegans in the United Kingdom: findings from the Feeding the Future (FEED) study

**DOI:** 10.64898/2026.07.09.26357676

**Authors:** William Bell, Izabella Lawson, Christopher Maronga, Sarah Clark, Kezia Gaitskell, Ben Lacey, Timothy J. Key, Keren Papier

**Affiliations:** Cancer Epidemiology Unit, Nuffield Department of Population Health, University of Oxford, Oxford OX3 7LF, UK; Clinical Trial Service Unit, Nuffield Department of Population Health, University of Oxford, Oxford; Nuffield Department of Population Health, University of Oxford, Oxford, OX3 7LF, UK

**Keywords:** Plant-Based, Biomarkers, Lipids, Micronutrient, Diet, Vegan

## Abstract

**Background & Aims:** The adoption of plant-based diets in the United Kingdom (UK) is increasing, which has potential health benefits, but may increase risk of inadequate intakes of some nutrients. We aimed to assess differences in biomarkers of nutritional status and disease across diet groups in UK adults.

**Methods:** This cross-sectional analysis included 124 omnivores, 131 flexitarians, 71 pescatarians, 124 vegetarians, and 183 vegans from the Feeding the Future (FEED) follow-up study (2024–2025). Capillary blood samples were collected and analysed for: haemoglobin; lipid measures (low- and high-density lipoprotein cholesterol (LDL-C and HDL-C), non-HDL-C, total cholesterol, and triglycerides); vitamins B12 and D. We estimated age- and sex-adjusted arithmetic or geometric mean concentrations and 95% confidence intervals of these biomarkers across diet groups. Participants taking lipid-lowering medications were excluded for analysis of lipid markers (n = 85).

**Results:** We observed differences in concentrations of cholesterol, triglycerides, and vitamin B12 across diet groups (P heterogeneity for all ≤ 0.03). Cholesterol markers (mmol/L) were lower with greater exclusion of animal foods (omnivores vs vegans, total cholesterol = −0.8; LDL-C = −0.7; HDL-C = −0.2). Triglyceride concentrations (mmol/L) were similar across groups, with slightly higher values in vegetarians (+0.2) and vegans (+0.1) compared with omnivores. Vitamin B12 concentrations (pmol/L) were highest in vegans and lowest in vegetarians compared to omnivores. Supplement users had higher vitamin B12 and D concentrations in all groups (P heterogeneity between strata = <0.001), while non-supplementing vegetarians and vegans had lower, but not deficient, vitamin B12 concentrations compared to omnivores (P heterogeneity between diet groups = <0.001).

**Conclusions:** In this contemporary UK cohort, those following plant-based diets had more favourable blood lipid profiles, with little evidence of vitamin B12 or D deficiency, or anaemia. Supplement use was associated with higher vitamin B12 and D concentrations, particularly among vegetarians and vegans.

## Introduction

Plant-based diets, in which individuals emphasise the consumption of plant foods, while limiting or completely excluding animal foods, are increasingly common in the United Kingdom (UK); data from the most recent National Diet and Nutrition Survey (NDNS) (2019-2023) suggest that approximately 3.5% of adults aged 19-64 identify as vegetarian (do not eat meat or fish) or vegan (do not consume any animal foods) (1), up from approximately 2% in 2008/2009 (2). Further, 4% report infrequent meat consumption, often described as a flexitarian dietary pattern. This shift has the potential to reduce the environmental impacts associated with high consumption of meat and dairy products (3) while also offering benefits for human health (4). Plant-based diets are typically characterised by higher intakes of dietary fibre and lower intakes of saturated fat (5). However, some plant-based diets, particularly vegan diets which are void of animal foods, may increase the risk of inadequate intakes of certain nutrients, including bioavailable iron, vitamin B12, vitamin D, and calcium, raising concerns about nutritional adequacy (5, 6).

Epidemiological evidence shows marked differences in nutrient intakes between meat-eaters and non-meat-eating vegetarians and vegans (7), which may influence nutritional status and disease risk. Previously, studies in western populations reported that vegetarians and vegans had a lower body mass index (BMI) as well as total cholesterol, and low density lipoprotein cholesterol (LDL-C) levels (8) but were also more likely to have lower concentrations of vitamins B12, and possibly vitamin D (9–11). Previous studies have also reported lower ferritin levels in vegetarians, and a slightly higher prevalence of anaemia in low and non-meat eaters (12, 13). In contrast, smaller, more contemporary studies between 2022 and 2024, typically including ∼100 vegetarians and vegans or fewer, have reported that as well as favourable lipid and anthropometric profiles, vegans had nutrient status often comparable to omnivores, possibly reflecting greater supplement use (14–16) or the wider availability of fortified plant-based alternatives in the modern food environment (17).

There is limited research on nutritional status and disease-related biomarker concentrations in adults consuming plant-based diets in contemporary settings, despite increasing adoption of these diets and the rapid expansion of plant-based meat and dairy alternatives (18). We therefore assessed these outcomes in UK adults consuming omnivorous, flexitarian, pescatarian, vegetarian, and vegan diets in the largest online cohort of UK adults consuming contemporary plant-based diets, the Feeding the Future (FEED) study.

## Methods

### Study Population and Description

Between 2022 and 2023, approximately 6,700 participants were recruited into FEED across the UK. Eligible participants were those aged 18 years and over, and recruitment was targeted at specific diet groups via various UK institutions, societies, and population groups including the Vegan Society, the Vegetarian Society, mainstream broadcasters, and food distributors to maximise uptake. Full details on the recruitment process have been described elsewhere (19). Participants joined the study by completing a 20-minute online questionnaire which collected sociodemographic, health, and lifestyle data, as well as dietary data via a food frequency questionnaire (FFQ) and questions on diet group (19). The study was approved by the University of Oxford Medical Sciences Interdivisional Research Ethics Committee (MS IDREC) (R79226/RE001) on 22 January 2022 and all participants provided informed consent. Participants who answered that they were happy to be contacted for future research projects and provided an email address in the baseline survey were eligible to take part in the follow-up survey. Between the 29^th^ of November 2024 and 30^th^ of August 2025 four invitation emails were sent approximately 2-3 months apart to all eligible participants with information on the follow-up survey and a link to a REDCap questionnaire, which included questions on diet group, an FFQ, health, medication and supplement use, and a consent form for taking part in the blood collection study. Participants who consented to take part in the blood collection were asked to provide their names and addresses for sending the capillary blood collection kits to their homes. Participants received a personalised report of their blood test results in return for taking part. This study is a cross sectional analysis of the blood biomarkers across self-reported diet groups using data from the follow-up assessment.

### Dietary Assessment

In the follow-up questionnaire, participants were asked to select a diet group that best described their current diet from the following options: omnivore (consume meat frequently), flexitarian (infrequent meat eating), pescatarian (consume fish but no meat), vegetarian (do not consume meat or fish), and vegan (do not consume any animal-source foods). More detailed dietary data were collected via the FFQ, which was an adapted version of the validated European Prospective Investigation into Cancer and Nutrition (EPIC) study FFQ, used in EPIC-Oxford, EPIC-Norfolk, Whitehall II, the Fenland Study, and the UK Women’s Cohort (20–22). For FEED, the FFQ was adapted to include a wider variety of contemporary foods such as plant-based meat and milk alternatives, as well as questions on protein supplement use and product fortification. Data on micronutrient supplement use was also added to the questionnaire, where participants were asked if they take supplements regularly, and if so to report the brand and quantity of any multivitamin and single nutrient supplements used. Portion size and quantitative intake estimates (in grams or millilitres) were estimated using standard portion sizes (23, 24) or using product manufacturers’ website information when these were not available. Nutrient intakes were calculated using the UK Nutrient Databank (NDB) extract version: UK_NDB_1.2, 2023 (25), and FoodDB (up to May 2022) (26), and checked against online retailers for plant-based alternatives not incorporated in the NDB (19).

### Blood Biomarkers

Blood samples were collected via at home finger prick capillary testing kits provided and processed by Thriva ltd (London, UK). Participants received a testing kit through the postal service which included instructions for sample collection and return to Thriva ltd, who analysed the samples. Participants were advised that two collection tubes were required to measure all of the biomarkers using ∼10-15 drops of blood microlitres into a collection tube containing anticoagulant (250 microliters for haemoglobin and 600 microliteres for the remaining biomarkers). Email reminders were sent out to participants who had not returned their samples from approximately 2 weeks after receiving the kit. Overall 6 biomarkers were assayed, and a further 4 calculated from these measurements. Serum vitamin B12 and vitamin D (measured as 25-hydroxyvitamin D_3_) were analysed via a Roche Cobas e801, using an electrochemiluminescence immunoassay. Haemoglobin was measured using a Beckman Coulter DxH 900 (before 24th April 2025) or a Horiba Yumizen 1500H / 550H (after 24th April 2025) using spectrophotometric testing. Serum total cholesterol, high density lipoprotein cholesterol (HDL-C), and triglyceride markers were analysed using a Roche Cobas c503, using an enzymatic colorimetric assay. Ratio markers, low density lipoprotein cholesterol (LDL-C), and non-HDL-C were calculated from these parameters.

### Covariates

Covariates used in this analysis were as follows: age, sex, height and weight used to calculate body mass index (BMI) (weight (kg)/height (m)^2^), smoking status, alcohol intake, supplement use, season of blood collection, and transit time. All covariates except height were self-reported and derived from the follow-up assessment questionnaire. Self-reported height was taken from the baseline assessment.

### Statistical Analysis

Descriptive characteristics and biomarker sample failure rates were analysed qualitatively using contingency tables to examine proportions (n (%)) in the total analysis sample and across diet groups. Multivariable linear regression models were used to investigate the associations between self-reported diet group and biomarker concentrations, with adjusted means calculated for each respective diet group using the least squares mean statistic via the emmeans package (27). For all analyses, omnivores were set as the reference category. Three models are presented. Model 1 was adjusted for age (continuous: years), and sex (categorical: women, men). Model 2 was adjusted for Model 1 plus BMI (categorical: tertials), and Model 3 was adjusted for Model 2 plus smoking status (categorical: never, ever), and alcohol intake (continuous: g/day). Season of measurement was added to all models for vitamin D (categorical: April-September (non-Autumn and Winter months), October-March (Autumn and Winter months)), defined based on NHS guidance as months where people are recommended to supplement (28). Model 1 was considered the primary model, as the small sample size and relative homogeneity of the cohort reduced the robustness of estimates from models adjusted for additional covariates. As such, haemoglobin is also presented using Model 1 only due to a greater sample failure rate. For analysis of blood lipids, participants taking lipid lowering medications (cholesterol lowering medications including statins) were excluded (n = 85). Vitamin B12, vitamin D, and triglycerides were log transformed to approximate a normal distribution, therefore adjusted means represent the geometric mean for these biomarkers. We additionally used established clinical reference ranges for biomarkers derived from the National Institute for Health and Care Excellence (NICE) and the National Health Service (NHS) guidance (29–32) to assess the proportion of participants within these ranges using unadjusted biomarker concentrations (presented as n (%) of diet group). The reference ranges used for classification are defined in the respective results table and text.

We conducted sensitivity analyses for biomarkers where vitamin and mineral supplementation were relevant, namely vitamin B12, vitamin D, and haemoglobin, stratifying analyses by vitamin B12, vitamin D, and iron supplement use (users and non-users) respectively. Supplement use was defined as either use of a multivitamin and mineral supplement or a vitamin B12, vitamin D, or iron specific supplement (multivitamins excluding minerals were not counted as iron supplementation). Additionally, those taking medically prescribed vitamin D were also considered supplement users (n = 17). Due to the low number of non-white participants in the cohort, we could not perform a stratified analysis by ethnic group and instead conducted a sensitivity analysis excluding non-white participants (n = 25). Further, we also conducted a sensitivity analysis adjusting for sample transit time (continuous: hours) due to the potential impact of transit time on sample stability. Transit time was calculated as the difference between the time of sample collection and time of sample analysis, and was log transformed to approximate a normal distribution, while 2 people with missing collection time data were excluded. P values for significant between group differences, and between strata of supplement use (P for heterogeneity) were obtained from multivariate ANCOVA. Where the global P for heterogeneity between groups was statistically significant (P <0.05), we conducted pairwise comparisons using Wald’s t-tests, adjusted for multiple testing using a Bonferroni correction. Compact letter display was generated to indicate pairwise statistical significance using the multicomp package (33). All analyses were conducted in R (version 4.1.0) (34).

## Results

### Recruitment and Sample Availability

A total of 3,590 invitation emails were sent out inviting participants to take part in the follow-up survey. Of these, 1,078 participants agreed to take part in the blood collection, of whom 758 returned a completed test kit to Thriva. From those who returned their kit, 89 had a nonviable sample, leaving 669 participants with at least one viable biomarker for analysis. Exclusions for the study were as follows: pregnancy (n = 5), and missing covariate data (n = 31), leaving 633 participants eligible for inclusion. There were 124 omnivores, 131 flexitarians, 71 pescatarians, 124 vegetarians, and 183 vegans included in the maximal analysis sample. A full flow chart of the analysis sample can be found in **Supplementary Figure S1**. The mean (SD) transit time in the post for samples was 2.1 (±1.3) days (median 1.7 days, range 0.7-14.5) and all but 3 of the participants blood samples were analysed within 7 days of sample collection. The total number of available samples for each specific biomarker is presented in **Supplementary Table S1** including the reasons for void or incomplete samples. Of the 758 participants who returned a sample, approximately 80% of samples were successfully tested for at least one biomarker with the exception of haemoglobin due to a large proportion of clotted samples (33.4%) and samples with insufficient blood (29.6%). The main reasons for void or incomplete samples for the other biomarkers were haemolysis (range across biomarkers: 0.4-9.5%), insufficient volume (range across biomarkers: 3.4-29.6%), and one of the samples not returned to Thriva ltd. (1.5% across all biomarkers). Other reasons included unlabelled samples, contamination during self-sampling (e.g. through EDTA contamination if blood was swapped between the two collection tubes in the kit), mismatched samples, and sample leakage although these were generally <1% across biomarkers.

### Descriptive Characteristics of the Cohort

Descriptive characteristics of the full study population are summarised in **Table 1**, and those of the cohort for lipid markers analyses (excluding 85 participants on lipid lowering medications) are presented in **Supplementary Table S2.** The mean (±SD) age of all participants was 58.1 (±13.2) years; vegetarians were oldest on average (61.5 (±11.5) years) and vegans the youngest (54.1 (±13.4) years). Compared with omnivores (25.7 (±6.8) kg/m^2^), the mean (±SD) BMI was lower in all other diet groups, lowest in vegans (23.7 (±3.9) kg/m^2^) and flexitarians (23.7 (±6.3) kg/m^2^). Total energy intake was broadly comparable across groups, while alcohol intake was slightly higher among omnivores and vegetarians (9.9 g/day) than other diet groups (<8.5g/day). Pescatarians had the highest prevalence of ever smoking (40.8%), and flexitarians the lowest (30.5%). Use of lipid-lowering medications was most common in pescatarians (18.3%) and least common in vegans (8.7%). Supplement use was generally high among all diet groups (>70%) but higher in all other diet groups compared with omnivores, with the highest usage (89.6%) observed among vegans. The majority (83.7%) of participants in all diet groups reported long-term dietary adherence (≥5 years), with the highest adherence observed among vegetarians (96.8%). Characteristics and trends in the subset excluding lipid lowering medications were similar to the full study population.

**Table 1:**
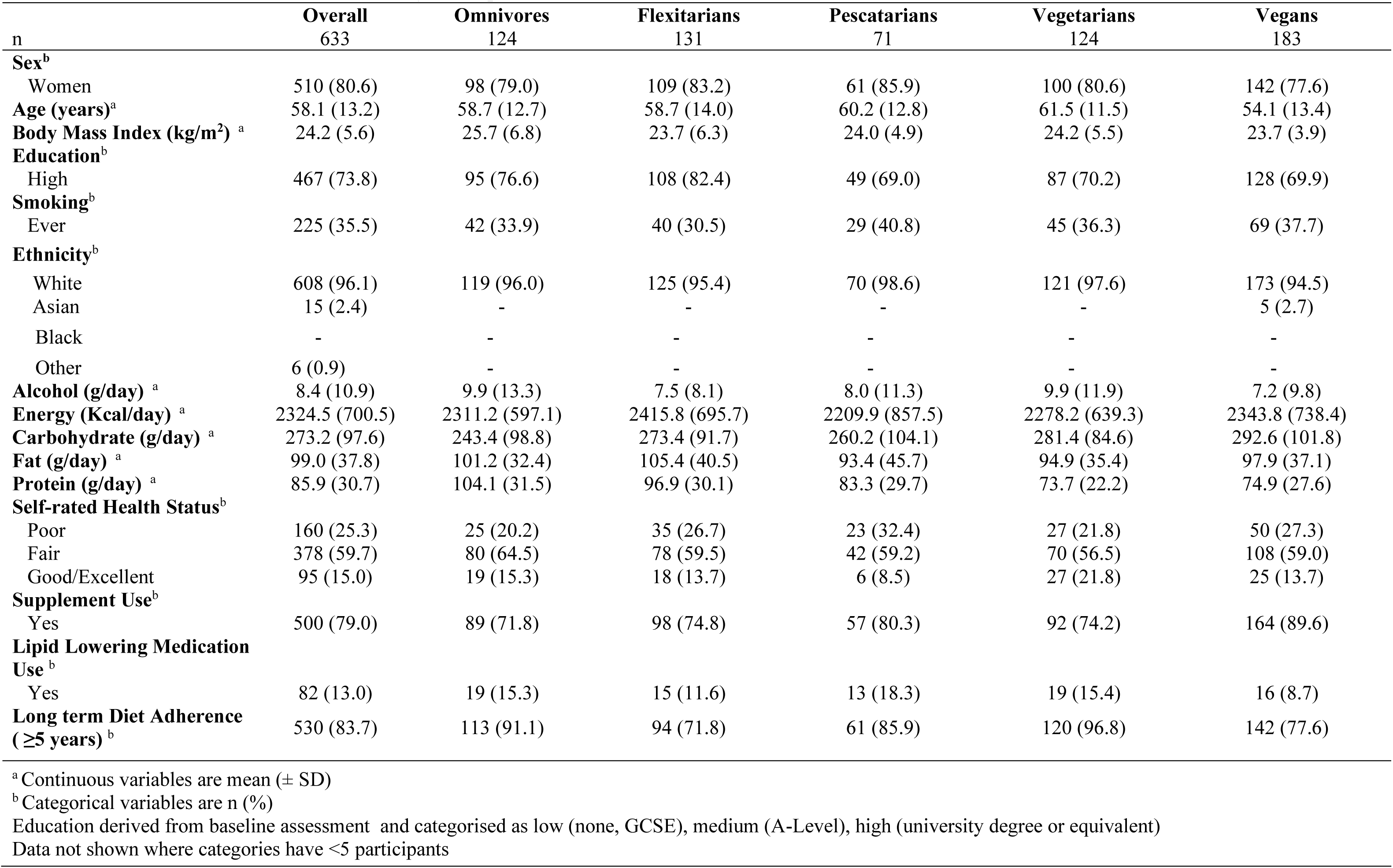
Cohort characteristics of the full analysis sample (N = 633)

|  | Overall | Omnivores | Flexitarians | Pescatarians | Vegetarians | Vegans |
| --- | --- | --- | --- | --- | --- | --- |
| n | 633 | 124 | 131 | 71 | 124 | 183 |
| **Sex<sup>b</sup>** |  |  |  |  |  |  |
| Women | 510 (80.6) | 98 (79.0) | 109 (83.2) | 61 (85.9) | 100 (80.6) | 142 (77.6) |
| **Age (years)<sup>a</sup>** | 58.1 (13.2) | 58.7 (12.7) | 58.7 (14.0) | 60.2 (12.8) | 61.5 (11.5) | 54.1 (13.4) |
| **Body Mass Index (kg/m<sup>2</sup>)<sup>a</sup>** | 24.2 (5.6) | 25.7 (6.8) | 23.7 (6.3) | 24.0 (4.9) | 24.2 (5.5) | 23.7 (3.9) |
| **Education<sup>b</sup>** |  |  |  |  |  |  |
| High | 467 (73.8) | 95 (76.6) | 108 (82.4) | 49 (69.0) | 87 (70.2) | 128 (69.9) |
| **Smoking<sup>b</sup>** |  |  |  |  |  |  |
| Ever | 225 (35.5) | 42 (33.9) | 40 (30.5) | 29 (40.8) | 45 (36.3) | 69 (37.7) |
| **Ethnicity<sup>b</sup>** |  |  |  |  |  |  |
| White | 608 (96.1) | 119 (96.0) | 125 (95.4) | 70 (98.6) | 121 (97.6) | 173 (94.5) |
| Asian | 15 (2.4) | - | - | - | - | 5 (2.7) |
| Black | - | - | - | - | - | - |
| Other | 6 (0.9) | - | - | - | - | - |
| **Alcohol (g/day)<sup>a</sup>** | 8.4 (10.9) | 9.9 (13.3) | 7.5 (8.1) | 8.0 (11.3) | 9.9 (11.9) | 7.2 (9.8) |
| **Energy (Kcal/day)<sup>a</sup>** | 2324.5 (700.5) | 2311.2 (597.1) | 2415.8 (695.7) | 2209.9 (857.5) | 2278.2 (639.3) | 2343.8 (738.4) |
| **Carbohydrate (g/day)<sup>a</sup>** | 273.2 (97.6) | 243.4 (98.8) | 273.4 (91.7) | 260.2 (104.1) | 281.4 (84.6) | 292.6 (101.8) |
| **Fat (g/day)<sup>a</sup>** | 99.0 (37.8) | 101.2 (32.4) | 105.4 (40.5) | 93.4 (45.7) | 94.9 (35.4) | 97.9 (37.1) |
| **Protein (g/day)<sup>a</sup>** | 85.9 (30.7) | 104.1 (31.5) | 96.9 (30.1) | 83.3 (29.7) | 73.7 (22.2) | 74.9 (27.6) |
| **Self-rated Health Status<sup>b</sup>** |  |  |  |  |  |  |
| Poor | 160 (25.3) | 25 (20.2) | 35 (26.7) | 23 (32.4) | 27 (21.8) | 50 (27.3) |
| Fair | 378 (59.7) | 80 (64.5) | 78 (59.5) | 42 (59.2) | 70 (56.5) | 108 (59.0) |
| Good/Excellent | 95 (15.0) | 19 (15.3) | 18 (13.7) | 6 (8.5) | 27 (21.8) | 25 (13.7) |
| **Supplement Use<sup>b</sup>** |  |  |  |  |  |  |
| Yes | 500 (79.0) | 89 (71.8) | 98 (74.8) | 57 (80.3) | 92 (74.2) | 164 (89.6) |
| **Lipid Lowering Medication Use<sup>b</sup>** |  |  |  |  |  |  |
| Yes | 82 (13.0) | 19 (15.3) | 15 (11.6) | 13 (18.3) | 19 (15.4) | 16 (8.7) |
| **Long term Diet Adherence (≥5 years)<sup>b</sup>** | 530 (83.7) | 113 (91.1) | 94 (71.8) | 61 (85.9) | 120 (96.8) | 142 (77.6) |

<sup>a</sup> Continuous variables are mean (± SD)
<sup>b</sup> Categorical variables are n (%)
Education derived from baseline assessment and categorised as low (none, GCSE), medium (A-Level), high (university degree or equivalent)
Data not shown where categories have <5 participants

### Biomarker Concentrations by Diet Group

Age and sex adjusted mean biomarker concentrations by diet group are shown in **Figure 1, Supplementary Figure S2,** and **Supplementary Table S3**. Compared with omnivores (339.2 pmol/L; 95% CI: 310.2-371.0), adjusted geometric mean concentrations of vitamin B12 were lower in vegetarians (302.1 pmol/L; 95% CI: 275.0-332.0), and flexitarians (323.7 pmol/L; 95% CI: 295.8 - 354.2), while pescatarians and vegans had 9 and 36 pmol/L higher vitamin B12 concentrations, respectively (P heterogeneity between diet groups = 0.01; pairwise significance for vegetarian’s vs vegans). Further adjustment for BMI, smoking and alcohol did not measurably change the associations, however the differences between flexitarians and vegans also became statistically significant in pairwise comparisons. Differences across diet groups for vitamin D (adjusted geometric mean) and haemoglobin (adjusted arithmetic mean) concentrations were not statistically significant (P heterogeneity between diet groups = 0.20 and 0.82 respectively) and this did not change with further adjustment for BMI, smoking, and alcohol intake.

**Figure 1:**
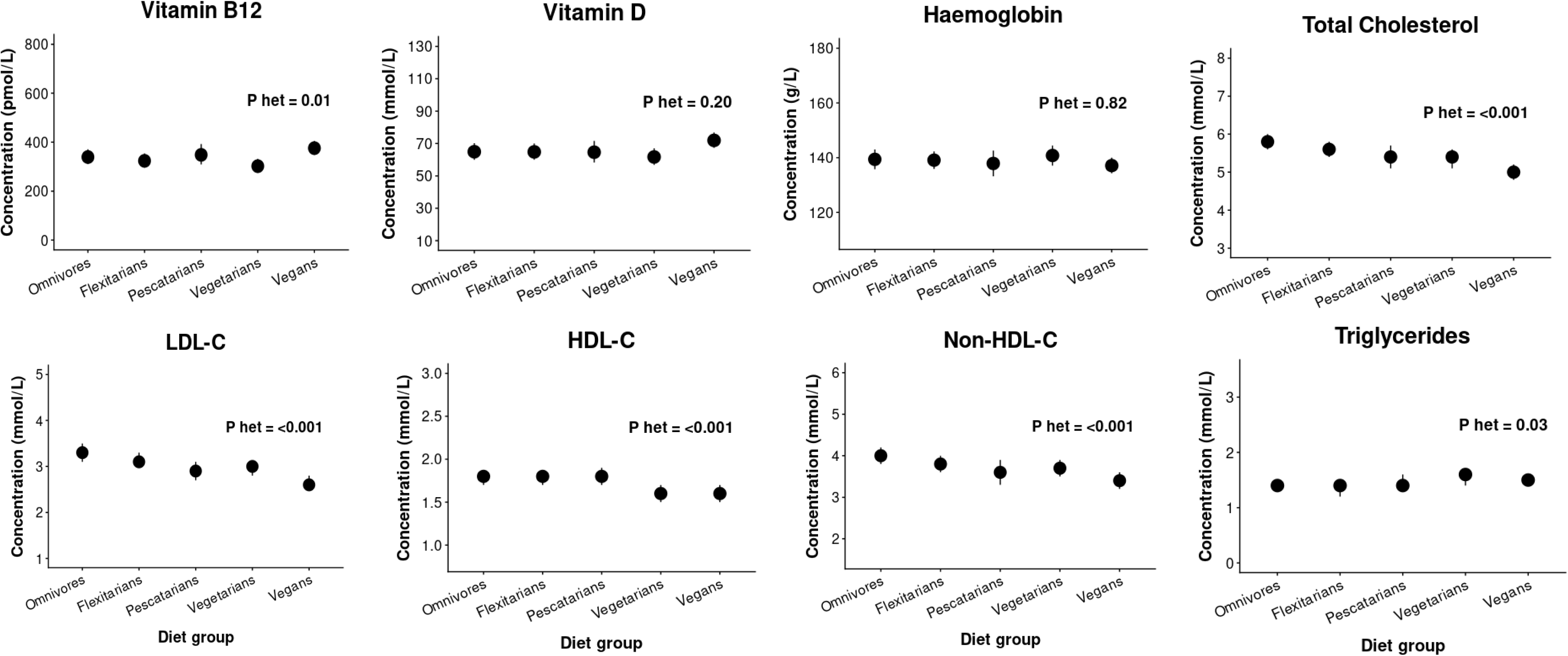
Age and sex adjusted concentrations of nutritional status and health-related biomarkers across diet groups in FEED. Values are adjusted arithmetic mean values (95% Confidence Intervals) and P heterogeneity (P het) for between group differences. Adjusted geometric means are presented for vitamin B12, D, and triglycerides. Model adjusted for age (years) and sex (women, men). Vitamin D additionally adjusted for season of measurement (October-March, April-September) The number of participants per group for each biomarker can be found in **Supplementary Table S3** Participants taking lipid lowering medications (n = 85) excluded for lipid markers HDL-C = High-Density Lipoprotein Cholesterol; LDL-C = Low-Density Lipoprotein Cholesterol.; P het = P for heterogeneity.

Compared with omnivores (5.8 mmol/L; 95% CI: 5.6- 6.0), adjusted arithmetic mean concentrations of total cholesterol decreased with greater animal food exclusion, and was lowest in vegans (5.0 mmol/L; 95% CI: 4.8-5.2) (P heterogeneity between diet groups = <0.001; pairwise significance for omnivores vs vegetarians, omnivores vs vegans, and flexitarians vs vegans). These associations were unchanged after adjustment for BMI, smoking and alcohol intake. Both LDL-C and non-HDL-C showed similar patterns. Adjusted arithmetic mean concentrations were highest in omnivores (LDL-C = 3.3 mmol/L; 95% CI: 3.1- 3.5, non-HDL-C = 4.0 mmol/L; 95% CI: 3.8- 4.2) and lowest in vegans (LDL-C = 2.6 mmol/L; 95% CI: 2.5- 2.8, non-HDL-C = 3.4 mmol/L; 95% CI: 3.2- 3.6) (P heterogeneity between diet groups for LDL-C and non-HDL-C both <0.001; pairwise significance: LDL-C = omnivores vs all other diet groups except flexitarians, flexitarians vs vegans, and vegetarians vs vegans; non-HDL = omnivores vs vegans and flexitarians vs vegans). Following adjustment for smoking, alcohol intake and BMI, pairwise significance in LDL-C concentrations between omnivores and vegans, flexitarians and vegans, and vegetarians and vegans remained, and non-HDL-C was unaffected. Adjusted arithmetic mean concentrations of HDL-C were similar in omnivores, flexitarians and pescatarians (all approximately 1.8 mmol/L), but lower in vegetarians and vegans (both 1.6 mmol/L; P heterogeneity between diet groups = <0.001; pairwise significance for vegans vs omnivores, flexitarians, and pescatarians). After adjustment, differences between omnivores and vegetarians were also statistically significant. Adjusted geometric mean triglyceride concentrations differed modestly across groups being similar in omnivores, flexitarians and pescatarians (∼1.4 mmol/L) and slightly higher in vegetarians and vegans (1.5-1.6 mmol/L; P heterogeneity between diet groups = 0.03; no pairwise significance). Further adjustment did not materially affect these findings. Compared with omnivores (3.6; 95% CI: 3.4-3.8) the adjusted arithmetic mean ratio of total cholesterol to HDL-C was lower in all non-omnivorous groups (range 3.2-3.5) (P heterogeneity between diet groups = 0.009; no pairwise significance) while the triglyceride-to-HDL-C ratio did not differ across diet groups (P heterogeneity between diet groups = 0.30), and results were unchanged after adjustment.

### Proportion of Cohort Out of Range

Classification of unadjusted biomarker concentrations into venous based clinical reference range categories are presented in **Table 2**. Overall, the majority of all diet groups were not deficient in vitamin B12 (≥133pmol/L), ranging from 94.8% in vegetarians to 98.4% in omnivores and flexitarians. Similarly, the majority of participants in all diet groups were not deficient in vitamin D (≥25nmol/L), ranging from 95.5% in vegetarians to 100% in flexitarians, and were also not anaemic (≥130 g/L for men and ≥115 g/L for women), ranging from 90.2% in omnivores, to 97.1% in vegans. A substantial portion of participants in all diet groups were above the recommended reference range for total cholesterol (≥5 mmol/L); prevalence ranged from 63.8%-79.0% in omnivores, flexitarians, pescatarians, and vegetarians, and was notably lower in vegans (46.8%). LDL-C and non-HDL-C followed a similar pattern, whereby the prevalence of high LDL-C (≥3 mmol/L) and non-HDL-C (≥4 mmol/L) were higher in omnivores, flexitarians, and vegetarians (LDL-C = 52.7-65.7%, non-HDL-C = 41.9 -47.0%), slightly lower in pescatarians (LDL-C = 44.8%, non-HDL-C = 34.5%), and lowest in vegans (LDL-C = 29.7%, non-HDL-C = 24.1%). The prevalence of low HDL-C levels (<1 mmol/L in men and <1.2 mmol/L in women) was highest among omnivores, vegetarians and vegans (>7%), and the prevalence of hypertriglyceridemia (≥2.3 mmol/L) was highest among vegetarians (18.3%), followed by vegans (13.9%), and lower in pescatarians, flexitarians, and omnivores (range: 9.0-12.1%).

**Table 2:** Prevalence of participants in each diet group across biomarker reference ranges in FEED.

|  |  |  |  |  |  |
| --- | --- | --- | --- | --- | --- |
|  | Omnivores | Flexitarians | Pescatarians | Vegetarians | Vegans |
| **Vitamin B12 (n = 616)** |  |  |  |  |  |
| Not Deficient ($\geq 133$ pmol/ L) | 122 (98.4) | 126 (98.4) | 67 (97.1) | 110 (94.8) | 174 (97.2) |
| **Vitamin D (n = 590)** |  |  |  |  |  |
| Not Deficient ($\geq 25$ nmol/L) | 112 (99.1) | 126 (100) | 66 (98.5) | 106 (95.5) | 170 (98.3) |
| **Haemoglobin (n = 230)** |  |  |  |  |  |
| Not Anaemic ($\geq 130$ g/L for men and $\geq 115$ g/L for women) | 37 (90.2) | 53 (96.4) | 21 (91.3) | 38 (92.7) | 68 (97.1) |
| **Total cholesterol (n = 519)** |  |  |  |  |  |
| High ($\geq 5$ mmol/L) | 79 (79.0) | 85 (77.3) | 37 (63.8) | 65 (69.9) | 74 (46.8) |
| **HDL-C (n = 541)** |  |  |  |  |  |
| Low ($< 1$ mmol/L in men and $< 1.2$ mmol/L in women) | 8 (7.6) | - | - | 8 (8.1) | 12 (7.2) |
| **LDL-C (n = 513)** |  |  |  |  |  |
| High ($\geq 3$ mmol/L) | 65 (65.7) | 65 (59.1) | 26 (44.8) | 48 (52.7) | 46 (29.7) |
| **Non-HDL-C (n = 519)** |  |  |  |  |  |
| High ($\geq 4$ mmol/L) | 47 (47.0) | 47 (42.7) | 20 (34.5) | 39 (41.9) | 38 (24.1) |
| **Triglycerides (n = 519)** |  |  |  |  |  |
| High ($\geq 2.3$ mmol/L) | 8 (8.0) | 13 (11.8) | 7 (12.1) | 17 (18.3) | 22 (13.9) |
| Values are n (%) |  |  |  |  |  |
| Biomarker concentrations used for categorisation are unadjusted |  |  |  |  |  |
| Reference ranges derived from National Institute of Health and Care Excellence guidance and National Health Service Guidelines |  |  |  |  |  |
| Data not shown where categories have $< 5$ participants | | | | | |
| Participants taking lipid lowering medications (n = 85) excluded for lipid markers |  |  |  |  |  |
| HDL = High-Density Lipoprotein cholesterol; LDL = Low-Density Lipoprotein cholesterol |  |  |  |  |  |

### Sensitivity Analysis

In analyses stratified by supplement use, vitamin B12 and D concentrations were higher in supplement users, whereas haemoglobin concentrations did not differ (P heterogeneity between strata: vitamin B12 = <0.001, vitamin D = <0.001, haemoglobin = 0.11) (**Figure 2**). Across these biomarkers, we observed significant between group differences in non-supplement users for vitamin B12 only (P heterogeneity between diet groups = <0.001). In non-supplement users, compared to omnivores (313.0 pmol/L; 95% CI: 285.6-343.0), adjusted geometric mean concentrations were lower in flexitarians (290.1 pmol/L; 95% CI: 262.8-320.2), pescatarians (301.4 pmol/L; 95% CI: 262.4-346.1), vegetarians (226.8 pmol/L; 95% CI: 202.0- 254.5), and vegans (247.6 pmol/L; 95% CI: 213.5 - 287.2), with pairwise significance between vegetarians, and omnivores, flexitarians, and pescatarians. After further adjustment for BMI, smoking and alcohol intake, the difference between omnivores and vegans was also statistically significant (**Supplementary Table S4**). Excluding non-white participants and adjusting for sample transit time did not change the associations (**Supplementary Table S5 and S6**).

**Figure 2:**
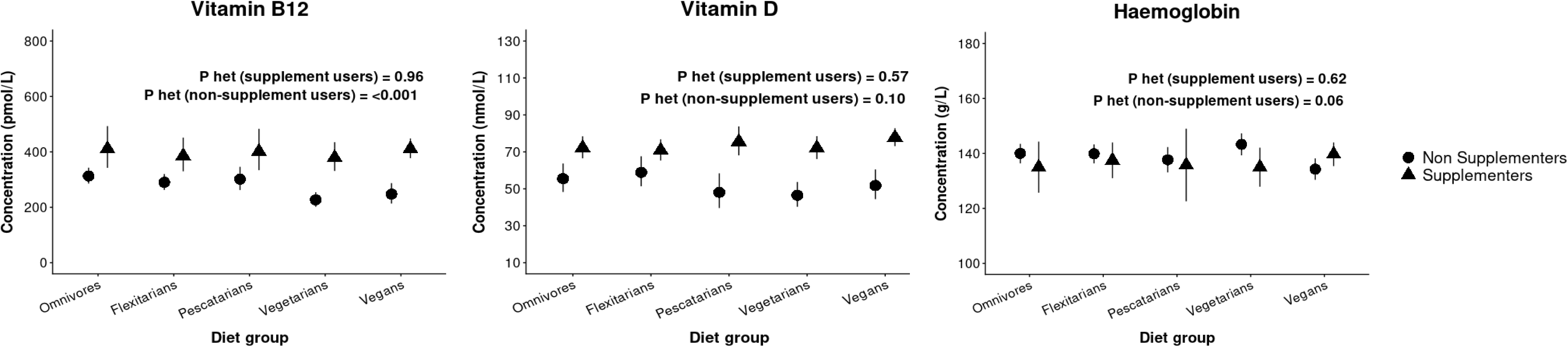
Age and sex adjusted concentrations of health-related biomarkers across diet groups in FEED stratified by supplementation use (vitamin B12, vitamin D, and iron respectively) Values are adjusted mean values (95% Confidence Intervals) and P heterogeneity (P het) for between group differences. Adjusted geometric means are presented for vitamin B12 and D, and adjusted arithmetic mean for haemoglobin. Model adjusted for age (years) and sex (women, men). Vitamin D additionally adjusted for season of measurement (October-March, April-September)). The number of participants per group for each biomarker can be found in **Supplementary Table S4** P het = P for heterogeneity.

## Discussion

In this contemporary cohort of UK adults, we observed differences in key nutrition and health related biomarker concentrations across diet groups. Notably, lipid markers were lower with greater exclusion of animal foods, with the exception of triglycerides which were marginally higher in vegetarians and vegans compared to other groups. In particular, differences between omnivores and vegans were largest for LDL-C, total cholesterol and Non-HDL-C, while differences in HDL-C cholesterol were smaller. Vitamin B12 concentrations differed by diet group, with vegans showing the highest levels and vegetarians the lowest compared with omnivores, while vitamin D and haemoglobin concentrations did not differ. Across all groups, non-supplement users had lower vitamin B12 and D concentrations than supplement users, but particularly for vitamin B12 among non-supplementing vegetarians and vegans when compared to omnivores, suggesting supplementation is contributing more to adequacy of vitamin B12 concentrations in these groups. Importantly even non-supplement users in all diet groups were on average not deficient in vitamins B12 and D, or anaemic, suggesting that in FEED contemporary plant-based diets were sufficient to avoid substantial deficiency of these nutrients.

Our findings for the lipid biomarkers are broadly consistent with smaller contemporary studies across Europe (14, 15), as well as older findings from large comparable cohorts (8, 35–38). These analyses similarly show a somewhat linear trend between greater animal food exclusion and lower total cholesterol, non-HDL-C, and LDL-C, with the magnitude of difference between omnivores and vegetarians and vegans ranging from approximately −0.50 to −1.20 mmol/L for total cholesterol, and approximately −0.10 to −1.10 mmol/L for non-HDL-C and LDL-C. Notably, similar to these analyses, we found minimal attenuation after adjustment for BMI, and also smoking and alcohol intake. We observed slightly lower HDL-C in vegetarians and vegans, which has also been reported previously (8, 35–37), and is likely a reflection of the lower total cholesterol in these participants (a finding also reflected in the lower total cholesterol:HDL-C ratio in vegans and vegetarians). Our lipid findings also align with meta-analyses of observational studies and randomised trials (RCTs) (39, 40). These meta-analyses of RCTs (n = 29 and 19 respectively) reported reductions of approximately −0.30 mmol/L in total and LDL-C following the implementation of vegan and vegetarian diets (pooled estimate), while differences between omnivores, vegetarians and vegans in cross-sectional observational studies (n = 30) were approximately −0.76 mmol/L for total cholesterol and −0.59 mmol/L for LDL-C. Consistent with our results, subgroup analysis also indicated effect sizes were greater for vegans than vegetarians. Differences in magnitude between within-group changes and between-group comparisons, as well as heterogeneity across study populations, are evident. However, the consistent inverse associations across both observational studies, with little impact of adjustment for potential confounders, and clinical trials strongly suggest a causal effect. This is likely, at least in part, related to a lower intake of saturated fat and greater intakes of polyunsaturated fat in plant based dietary patterns (5, 19). We observed a general trend towards marginally higher triglyceride levels in vegetarians and vegans, reported in observational studies but not clinical trials previously (39). Replacement of fat with carbohydrates may increase triglycerides (41, 42) which is also a feature of vegetarian and vegan diets in FEED, alongside lower fat and protein intake (19).

We observed differences in vitamin B12 concentrations across diet groups, although significant pairwise differences were limited to vegetarians and vegans. The prevalence of clinical deficiency was uncommon in all groups (<6%), but marginally higher in vegetarians than other groups, consistent with group means. Among supplement users, vitamin B12 concentrations did not differ by diet group, whereas among non-users, concentrations were lower in vegetarians and vegans, suggesting supplement use is important in these groups. These findings are broadly consistent with contemporary findings from Storz et al (14) where in a small cohort of 37 vegetarians and 38 vegans in Germany, vitamin B12 was highest in vegans and similar to those in omnivores, while being lowest in vegetarians, with differences in supplement use likely explaining the greater vitamin B12 concentrations in vegans, and lower concentrations in vegetarians. A recent meta-analysis of 19 observational studies assessing vitamin B12 status across different diet groups between 1999 and 2023, found that generally mean absolute vitamin B12 concentrations were not strongly indicative of deficiency in vegans (249 pmol/L) and vegetarians (252 pmol/L), but that these groups did have lower vitamin B12 concentrations than omnivores (385 pmol/L). Additionally, markers of functional vitamin B12 status such as holotranscobalamin, methylmalonic acid, and homocysteine indicated a greater risk of B12 insufficiency in these groups, particularly in non-supplementing vegans (10). With this said, absolute vitamin B12 concentrations in non-supplementing vegans in FEED are still considerably higher than those observed in non-supplementing vegans in the meta-analysis, and than those from the comparable EPIC Oxford cohort in the 1990s which suggests dietary intake of vitamin B12 is supporting vegans more compared to previous years, likely relating to the high consumption of commonly fortified plant milks and dairy alternatives in FEED (17, 19).

There was little difference in vitamin D status across diet groups, and deficiency was uncommon. Chan, Jaceldo-Siegl (43) observed similar findings when comparing vegetarians to omnivores in the Adventist Health Study-2 (AHS-2). However these observations are different from previous UK cohorts, and may reflect greater sun exposure related to geographical location and/or fortification and supplement use in AHS-2. Contrary to FEED, findings from EPIC-Oxford in the late 1990s showed vitamin D levels were significantly lower in vegans and vegetarians than in omnivores (11), which was subsequently also observed in the UK Biobank from data collected in the late 2000s (8). Notably, absolute concentrations in EPIC-Oxford were considerably higher than those in the UK Biobank however (and closer to those we observe in FEED), although this may also reflect methodological differences such as the assay used to analyse vitamin D. Stratification by vitamin D supplementation in FEED did result in lower concentrations across all diet groups, with absolute concentrations in non-supplement users similar to those observed in the UK Biobank. Thus differences in absolute concentrations between studies may also reflect levels of supplement use. Despite sensitivity analysis to exclude non-white participants, the smaller overall cohort size and targeted recruitment also limits analysis on ethnic differences which were reported on in prior studies.

Regarding haemoglobin, previous findings from the UK Biobank have observed a general trend towards lower haemoglobin levels and a greater prevalence of anaemia in vegans and vegetarians (12), although differences were generally not statistically significant. In FEED we did not observe this trend, and further we observe no between group differences across strata of iron supplementation, and a low prevalence of anaemia (<10%) for all groups. With this said, the number of samples available for haemoglobin in FEED was low as a result of issues with sample collection and reduces the comparability of these findings. In addition to this the older majority female sample in FEED limits investigation as to whether menopausal status may also modify these associations. Although nutrients other than iron, such as folate and zinc, may also impact haemoglobin levels, intakes in the FEED cohort were sufficient (19), and the similar haemoglobin levels and low prevalence of anaemia across diet groups do not suggest this is of concern.

This study is strengthened by the large number of vegetarians and vegan participants, the inclusion of pescatarians, and further the separation of flexitarians from high meat eaters compared to previous studies. This separation is likely to have reduced dilution of differences between groups, potentially allowing for clearer contrasts between diet groups. This study is also strengthened by the use of objective measurements of nutritional biomarker status across diet groups. Limitations of the study include the relatively moderate sample size of the overall cohort and targeted recruitment limiting large scale generalisability, also in relation to non-white populations in the UK given the study population was made up predominantly of white British participants. Further, the self-reported nature of the diet group classification may result in some misclassification bias, although previous publications in FEED show that intakes of key nutrients and foods in the FFQ are congruent with self-reported diet classifications (19). Comparability of capillary based blood samples to standard venous tests and sample instability due to transit time may be a source of inaccuracy in the study. However studies to date have shown acceptable correlations between the measurement methods (44–48), while sensitivity analysis adjusting for transit time did not change our findings. Additionally, as capillary samples were collected at home, samples are likely a mix of fasting and non-fasting measures, although the distribution of fasting and non-fasting samples across diet groups is likely to be random and thus unlikely to substantially bias estimates. We did not have available data to assess functional vitamin B12 status, which limits inference regarding vitamin B12 status beyond total vitamin B12. The majority of participants in this cohort used supplements, thus further contemporary studies are warranted in individuals that do not use supplements.

## Conclusions

In this large contemporary UK cohort, plant-based diets were associated with more favourable lipid profiles without evidence of substantial risk of deficiency in vitamin B12 or vitamin D, or risk of anaemia. The low prevalence of deficiency likely reflects both supplement use and/or the influence of food fortification. Supplement use was associated with higher vitamin B12 and D concentrations, but particularly for vitamin B12 in vegetarians and vegans. Future work should also investigate functional vitamin B12 status, and assess other nutrients of concern such as iodine, and the relative contributions of fortified foods and other dietary sources of iron, fat, vitamin B12, and vitamin D to nutritional status and disease biomarkers in adults consuming contemporary plant-based diets. Further studies are needed in populations with different fortification practices, supplement use, and sociodemographic characteristics.

## Acknowledgments

We would like to thank the FEED participants for taking the time to complete the survey.

## Author Contributions

W.B., T.J.K. and K.P. devised the study. W.B. and K.P. analysed the data and wrote the paper. I.L, C.M, and K.P conducted data collection. All authors helped with the interpretation of the findings. K.P. conceived and developed the cohort. All authors have read and agreed to the published version of the manuscript.

## Institutional Review Board Statement

This study was approved by the University of Oxford Medical Sciences Interdivisional Research Ethics Committee (MS IDREC) (R79226/RE001) on 22 January 2022.

## Informed Consent Statement

Informed consent was obtained from all subjects involved in the study.

## Data Availability Statement

The data access policy for the FEED study is available via the study website: https://www.ceu.ox.ac.uk/research/feeding-the-future-study-feed.

## Declaration of generative AI in scientific writing

During the preparation of this work the author(s) used ChatGPT to proof read and improve readability of text. After using this tool/service, the author(s) reviewed and edited the content as needed and take(s) full responsibility for the content of the publication.

