## Supplementary Material for "Comparison of selected nutritional status and disease biomarkers in omnivores, flexitarians, pescatarians, vegetarians, and vegans in the United Kingdom: findings from the Feeding the Future (FEED) study"

| | | |
|---|---|---|
| **Supplementary Figure S1** | Participant flow chart for the study………………………………………………. | Page 3 |
| **Supplementary Table S1** | Lab failure rates for biomarker testing across biomarkers……………………….. | Page 4 |
| **Supplementary Table S2** | Cohort characteristics of the lipid analysis subset……………………………….. | Page 4 |
| **Supplementary Figure S2** | Adjusted mean biomarker concentrations for lipid ratios by diet group……….. | Page 5 |
| **Supplementary Table S3** | Biomarker concentrations across diet groups in FEED by levels of adjustment for potential confounders……………………………………………………… | Page 6 |
| **Supplementary Table S4** | Sensitivity analysis of adjusted biomarker concentrations across diet groups stratified by supplementation……………………………………………………….. | Page 7 |
| **Supplementary Table S5** | Sensitivity analysis of adjusted biomarker concentrations across diet group excluding non-white participants………………………………………………… | Page 8 |
| **Supplementary Table S6** | Sensitivity analysis of adjusted biomarker concentrations across diet groups additionally adjusted for transit time…………………………………………... | Page 9 |

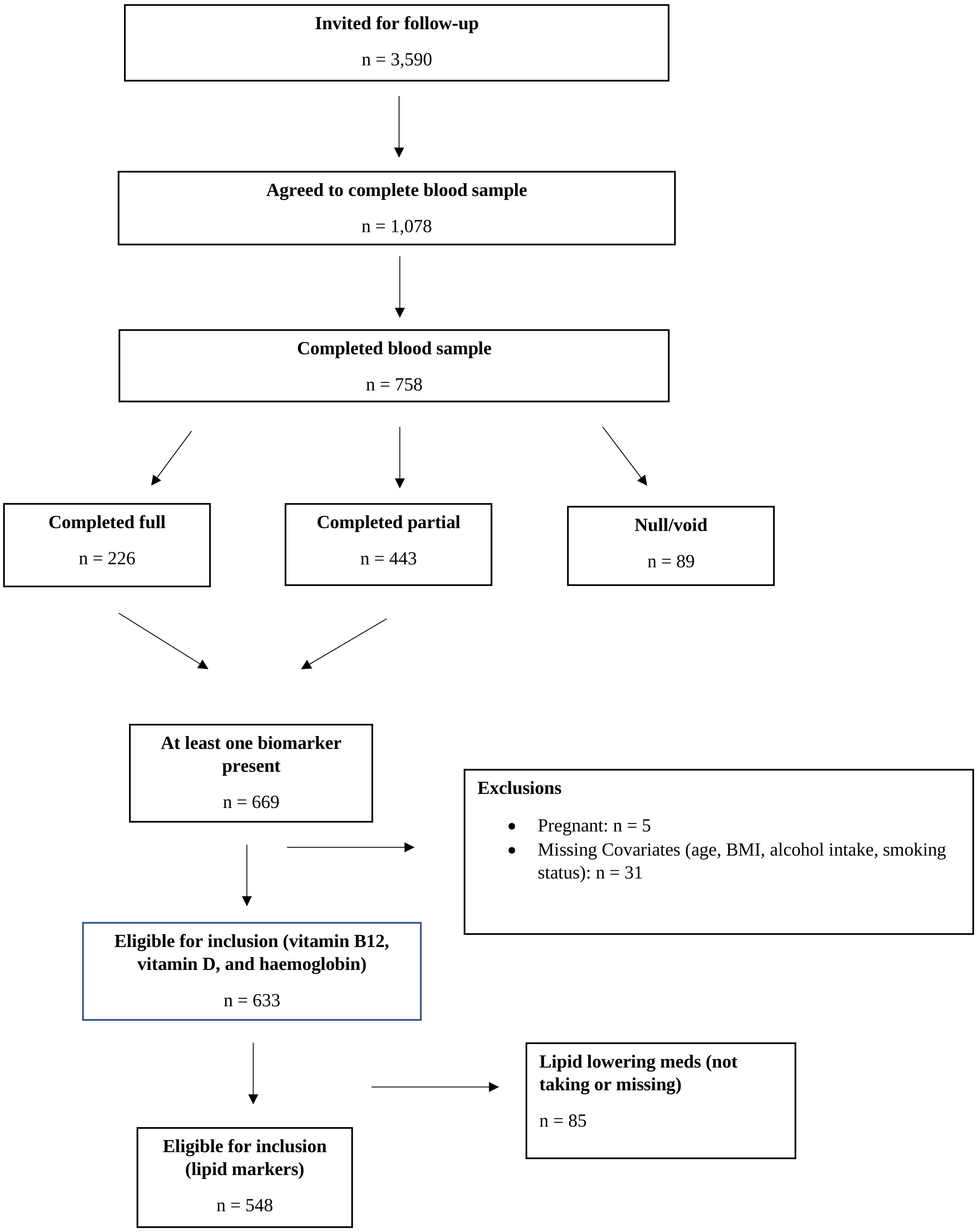


**Supplementary Figure S1: Participant flow chart for the study**

| Supplementary Table S1: Lab failure rates for biomarker testing across biomarkers | | | | | | |
|---|---|---|---|---|---|---|
| **Failure reason** | **Vitamin B12** | **Vitamin D** | **Haemoglobin** | **Total Cholesterol** | **HDL-C** | **Triglycerides** |
| Complete | 651 (85.9%) | 622 (82.1%) | 242 (31.9%) | 631 (83.2%) | 658 (86.8%) | 631 (83.2%) |
| Clotted | 0 (0.0%) | 0 (0.0%) | 253 (33.4%) | 0 (0.0%) | 0 (0.0%) | 0 (0.0%) |
| Contaminated | 6 (0.8%) | 5 (0.7%) | 3 (0.4%) | 6 (0.8%) | 6 (0.8%) | 6 (0.8%) |
| Delay | 2 (0.3%) | 2 (0.3%) | 2 (0.3%) | 2 (0.3%) | 2 (0.3%) | 2 (0.3%) |
| Haemolysed | 42 (5.5%) | 72 (9.5%) | 3 (0.4%) | 61 (8.0%) | 34 (4.5%) | 61 (8.0%) |
| Insufficient Volume | 26 (3.4%) | 26 (3.4%) | 224 (29.6%) | 27 (3.6%) | 27 (3.6%) | 27 (3.6%) |
| Mismatch | 5 (0.7%) | 5 (0.7%) | 5 (0.7%) | 5 (0.7%) | 5 (0.7%) | 5 (0.7%) |
| No sample received | 11 (1.5%) | 11 (1.5%) | 11 (1.5%) | 11 (1.5%) | 11 (1.5%) | 11 (1.5%) |
| Not tested | 6 (0.8%) | 6 (0.8%) | 6 (0.8%) | 6 (0.8%) | 6 (0.8%) | 6 (0.8%) |
| Sample leaked | 2 (0.3%) | 2 (0.3%) | 1 (0.1%) | 2 (0.3%) | 2 (0.3%) | 2 (0.3%) |
| Unlabelled | 7 (0.9%) | 7 (0.9%) | 8 (1.1%) | 7 (0.9%) | 7 (0.9%) | 7 (0.9%) |
| Total Failed | 107 (14.1) | 136 (17.9) | 516 (68.1) | 127 (16.8) | 100 (13.2) | 127 (16. 8) |

Data are number of samples (%)
All reported % are as a proportion of the total 758 test kits returned.
Note: Failure rates across calculated biomarkers (non-HDL-C, low density lipoprotein cholesterol, total cholesterol:HDL-C, and triglycerides:HDL-C match the maximal n of the total samples failed for their constituent biomarkers.
HDL-C = High Density Lipoprotein-Cholesterol

| Supplementary Table S2: Cohort characteristics of the lipid analysis subset | | | | | | |
|---|---|---|---|---|---|---|
| | **Overall** | **Omnivores** | **Flexitarians** | **Pescatarians** | **Vegetarians** | **Vegans** |
| n | 548 | 105 | 114 | 58 | 104 | 167 |
| **Sex**[a] | | | | | | |
| Women | 458 (83.6) | 86 (81.9) | 100 (87.7) | 52 (89.7) | 86 (82.7) | 134 (80.2) |
| **Age (years)** [b] | 56.6 (13.2) | 57.2 (12.8) | 57.3 (13.9) | 57.9 (12.8) | 60.5 (12.0) | 52.8 (13.1) |
| **Body Mass Index (Kg/m$^2$)** [b] | 24.0 (5.3) | 25.6 (7.0) | 23.5 (6.1) | 23.5 (4.1) | 23.7 (4.8) | 23.5 (3.9) |
| **Education**[a] | | | | | | |
| High | 407 (74.3) | 80 (76.2) | 93 (81.6) | 41 (70.7) | 76 (73.1) | 117 (70.1) |
| **Smoking**[a] | | | | | | |
| Ever | 191 (34.9) | 35 (33.3) | 38 (33.3) | 21 (36.2) | 34 (32.7) | 63 (37.7) |
| **Ethnicity**[a] | | | | | | |
| Asian | 13 ( 2.4) | - | - | - | - | 5 ( 3.0) |
| Black | - | - | - | - | - | - |
| Other | 6 ( 1.1) | - | - | - | - | - |
| White | 526 (96.0) | 102 (97.1) | 108 (94.7) | 57 (98.3) | 102 (98.1) | 157 (94.0) |
| **Alcohol (g/day)** [b] | 8.1 (10.5) | 9.9 (13.6) | 7.4 (8.0) | 7.5 (10.4) | 9.7 (11.4) | 6.7 (9.1) |
| **Energy (kcal/day)** [b] | 2313.1 (701.0) | 2289.1 (586.8) | 2397.5 (668.0) | 2191.7 (869.0) | 2266.9 (654.2) | 2341.6 (750.0) |
| **Carbohydrate (g/day)** [b] | 269.8 (98.3) | 236.6 (99.3) | 271.3 (89.6) | 256.3 (109.6) | 278.3 (82.9) | 289.0 (103.1) |
| **Fat (g/day)** [b] | 99.6 (37.8) | 101.7 (32.9) | 104.7 (39.6) | 93.8 (45.0) | 95.3 (36.5) | 99.4 (37.4) |
| **Protein (g/day)** [b] | 85.4 (31.0) | 103.8 (32.9) | 96.0 (28.8) | 82.4 (30.7) | 73.2 (23.1) | 75.3 (27.9) |
| **Health Status**[a] | | | | | | |
| Poor | 146 (26.6) | 23 (21.9) | 31 (27.2) | 20 (34.5) | 25 (24.0) | 47 (28.1) |
| Fair | 327 (59.7) | 68 (64.8) | 70 (61.4) | 34 (58.6) | 57 (54.8) | 98 (58.7) |
| Good/Excellent | 75 (13.7) | 14 (13.3) | 13 (11.4) | 4 (6.9) | 22 (21.2) | 22 (13.2) |
| **Supplement Use**[a] | | | | | | |
| Yes | 433 (79.0) | 77 (73.3) | 85 (74.6) | 46 (79.3) | 76 (73.1) | 149 (89.2) |
| **Long term Diet Adherence (≥5 years)** [a] | | | | | | |
| | 452 (82.5) | 95 (90.5) | 79 (69.3) | 48 (82.8) | 101 (97.1) | 129 (77.2) |

[a] Continuous variables are mean (± SD)
[b] Categorical variables are n (%)
Education derived from baseline assessment and categorised as low (none, GCSE), medium (A-Level), high (university degree or equivalent)
Data not shown where categories have <5 participants

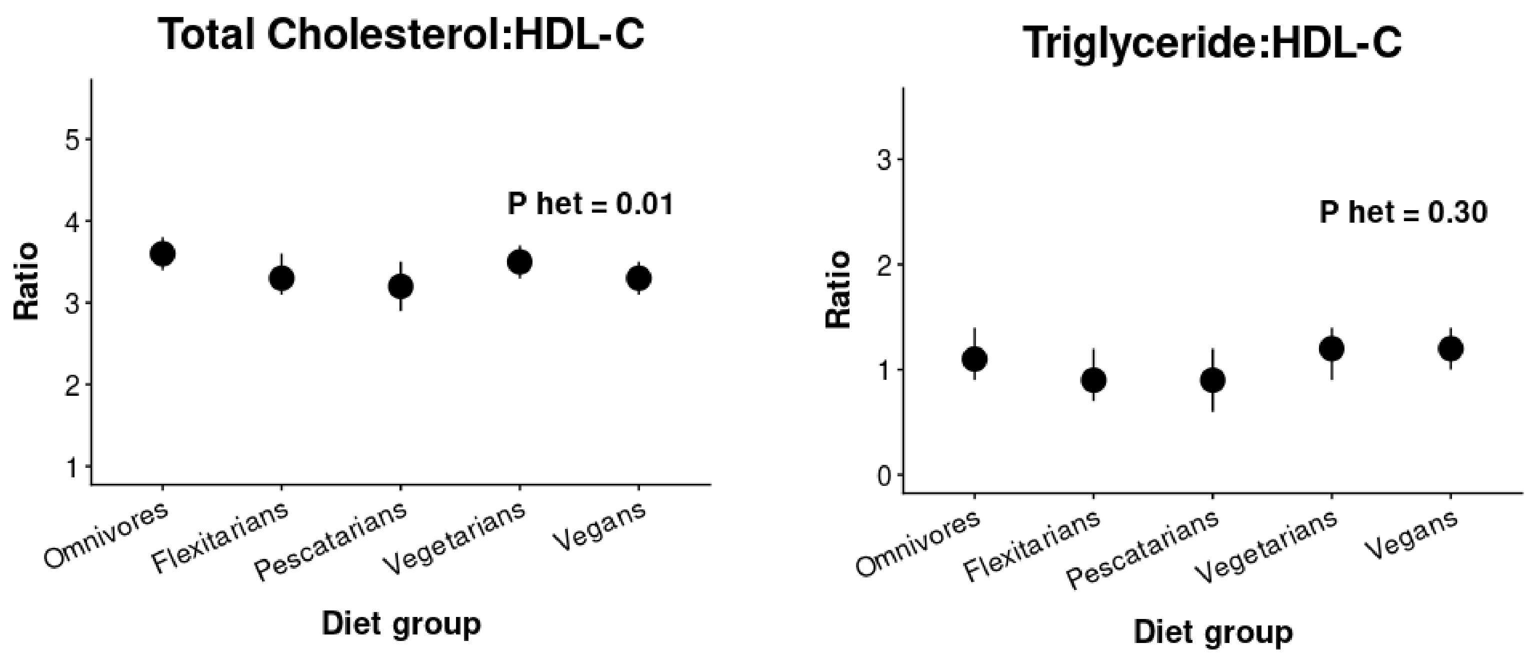


**Supplementary Figure S2: Adjusted mean biomarker concentrations for lipid ratios by diet group.**

Values are adjusted arithmetic mean values (95% confidence intervals)

Model adjusted for age (years) and sex (women, men).Vitamin D additionally adjusted for season of measurement (October-March, April-September)

The number of participants per group for each biomarker can be found in **Supplementary Table S3.**

Participants taking lipid lowering medications (n = 85) excluded.

HDL-C = High-Density Lipoprotein Cholesterol

**Supplementary Table S3: Biomarker concentrations across diet groups in FEED by levels of adjustment for potential confounders**

| Biomarker | | | Omnivorous | Flexitarians | Pescatarians | Vegetarians | Vegans | P for heterogeneity |
|---|---|---|---|---|---|---|---|---|
| **Vitamin B12 (pmol/L)** | **n** | **616** | **124** | **128** | **69** | **116** | **179** | |
| | Model 1 | | 339.2 (310.2, 371.0) a,b | 323.7 (295.8, 354.2) a,b | 348.2 (309.1, 392.3) a,b | 302.1 (275.0, 332.0) a | 375.6 (348.2, 405.2) b | 0.01 |
| | Model 2 | | 347.1 (317.5, 379.4) a,b | 320.9 (293.6, 350.7) a | 351.4 (312.4, 395.1) a,b | 305.3 (278.2, 335.0) a | 377.8 (350.6, 407.1) b | 0.008 |
| | Model 3 | | 353.2 (322.7, 386.6) a,b | 322.9 (295.3, 353.2) a | 354.0 (314.9, 398.0) a,b | 310.5 (282.7, 341.0) a | 379.8 (352.4, 409.3) b | 0.008 |
| **Vitamin D (nmol/L)** | **n** | **590** | **113** | **126** | **67** | **111** | **173** | |
| | Model 1 | | 64.9 (59.9, 70.2) | 64.8 (60.0, 70.0) | 64.6 (58.3, 71.6) | 61.7 (56.8, 67.0) | 71.9 (67.3, 76.8) | 0.20 |
| | Model 2 | | 65.6 (60.5, 71.2) | 65.0 (60.1, 70.3) | 65.3 (58.8, 72.6) | 62.1 (57.1, 67.5) | 72.3 (67.6, 77.4) | 0.20 |
| | Model 3 | | 65.7 (60.5, 71.4) | 65.4 (60.3, 70.8) | 65.5 (59.0, 72.9) | 62.2 (57.1, 67.7) | 72.5 (67.8, 77.6) | 0.20 |
| **Haemoglobin (g/L)** | **n** | **230** | **41** | **55** | **23** | **41** | **70** | |
| | Model 1 | | 139.4 (135.8, 143.0) | 139.1 (135.9, 142.3) | 137.9 (133.2, 142.6) | 140.8 (137.1, 144.4) | 137.1 (134.3, 140.0) | 0.82 |
| **Total Cholesterol (mmol/L)** | **n** | **519** | **100** | **110** | **58** | **93** | **158** | |
| | Model 1 | | 5.8 (5.6, 6.0) a | 5.6 (5.4, 5.8) a,b | 5.4 (5.1, 5.7) a,b,c | 5.4 (5.1, 5.6) b,c | 5.0 (4.8, 5.2) c | <0.001 |
| | Model 2 | | 5.8 (5.6, 6.0) a | 5.6 (5.4, 5.8) a,b | 5.4 (5.1, 5.7) a,b,c | 5.3 (5.1, 5.6) b,c | 5.0 (4.8, 5.2) c | <0.001 |
| | Model 3 | | 5.8 (5.5, 6.0) a | 5.6 (5.4, 5.8) a,b | 5.4 (5.1, 5.7) a,b,c | 5.3 (5.1, 5.6) b,c | 5.0 (4.8, 5.2) c | <0.001 |
| **LDL (mmol/L)** | **n** | **513** | **99** | **110** | **58** | **91** | **155** | |
| | Model 1 | | 3.3 (3.1, 3.5) a | 3.1 (3.0, 3.3) a,b | 2.9 (2.7, 3.1) b,c | 3.0 (2.8, 3.1) b | 2.6 (2.5, 2.8) c | <0.001 |
| | Model 2 | | 3.3 (3.1, 3.4) a | 3.2 (3.0, 3.3) a | 2.9 (2.7, 3.1) a,b | 2.9 (2.8, 3.1) a | 2.6 (2.5, 2.8) b | <0.001 |
| | Model 3 | | 3.3 (3.1, 3.4) a | 3.1 (3.0, 3.3) a | 2.9 (2.7, 3.1) a,b | 2.9 (2.7, 3.1) a | 2.6 (2.5, 2.8) b | <0.001 |
| **HDL (mmol/L)** | **n** | **541** | **105** | **113** | **58** | **99** | **166** | |
| | Model 1 | | 1.8 (1.7, 1.8) a | 1.8 (1.7, 1.8) a | 1.8 (1.7, 1.9) a | 1.6 (1.5, 1.7) a,b | 1.6 (1.5, 1.7) b | <0.001 |
| | Model 2 | | 1.8 (1.7, 1.9) a | 1.7 (1.7, 1.8) a,b | 1.8 (1.7, 1.9) a | 1.6 (1.5, 1.7) a,b | 1.6 (1.5, 1.7) b | <0.001 |
| | Model 3 | | 1.8 (1.7, 1.9) a | 1.7 (1.7, 1.8) a,b,c | 1.8 (1.7, 1.9) a,b | 1.6 (1.5, 1.7) b,c | 1.6 (1.5, 1.7) c | <0.001 |
| **Triglycerides (mmol/L)** | **n** | **519** | **100** | **110** | **58** | **93** | **158** | |
| | Model 1 | | 1.4 (1.3, 1.5) a | 1.4 (1.2, 1.5) a | 1.4 (1.3, 1.6) a | 1.6 (1.4, 1.7) a | 1.5 (1.4, 1.6) a | 0.03 |
| | Model 2 | | 1.3 (1.2, 1.5) a | 1.4 (1.3, 1.5) a | 1.4 (1.3, 1.6) a | 1.6 (1.4, 1.7) a | 1.5 (1.4, 1.6) a | 0.02 |
| | Model 3 | | 1.4 (1.2, 1.5) a | 1.4 (1.3, 1.5) a | 1.4 (1.3, 1.6) a | 1.6 (1.5, 1.8) a | 1.5 (1.4, 1.6) a | 0.02 |
| **Non-HDL Cholesterol (mmol/L)** | **n** | **519** | **100** | **110** | **58** | **93** | **158** | |
| | Model 1 | | 4.0 (3.8, 4.2) a | 3.8 (3.6, 4.0) a | 3.6 (3.3, 3.9) a,b | 3.7 (3.5, 3.9) a,b | 3.4 (3.2, 3.6) b | <0.001 |
| | Model 2 | | 4.0 (3.8, 4.2) a | 3.8 (3.6, 4.0) a | 3.6 (3.3, 3.8) a,b | 3.7 (3.5, 3.9) a,b | 3.4 (3.2, 3.5) b | <0.001 |
| | Model 3 | | 4.0 (3.8, 4.2) a | 3.8 (3.6, 4.0) a | 3.6 (3.3, 3.9) a,b | 3.7 (3.5, 3.9) a,b | 3.4 (3.2, 3.5) b | <0.001 |
| **Total Cholesterol:HDL Ratio** | **n** | **519** | **100** | **110** | **58** | **93** | **158** | |
| | Model 1 | | 3.6 (3.4, 3.8) a | 3.3 (3.1, 3.6) a | 3.2 (2.9, 3.5) a | 3.5 (3.3, 3.7) a | 3.3 (3.1, 3.5) a | 0.009 |
| | Model 2 | | 3.5 (3.3, 3.7) a | 3.4 (3.1, 3.6) a | 3.1 (2.9, 3.4) a | 3.5 (3.3, 3.7) a | 3.3 (3.1, 3.4) a | 0.005 |
| | Model 3 | | 3.6 (3.3, 3.8) a | 3.4 (3.2, 3.6) a | 3.2 (2.9, 3.4) a | 3.5 (3.3, 3.7) a | 3.3 (3.1, 3.4) a | 0.004 |
| **Triglyceride:HDL Ratio** | **n** | **519** | **100** | **110** | **58** | **93** | **158** | |
| | Model 1 | | 1.1 (0.9, 1.4) | 0.9 (0.7, 1.2) | 0.9 (0.6, 1.2) | 1.2 (0.9, 1.4) | 1.2 (1.0, 1.4) | 0.30 |
| | Model 2 | | 1.1 (0.8, 1.3) | 1.0 (0.7, 1.2) | 0.9 (0.6, 1.2) | 1.2 (0.9, 1.4) | 1.2 (1.0, 1.3) | 0.28 |
| | Model 3 | | 1.1 (0.9, 1.4) | 1.0 (0.8, 1.2) | 1.0 (0.7, 1.3) | 1.2 (1.0, 1.5) | 1.2 (1.0, 1.4) | 0.27 |

Values are adjusted arithmetic mean values (95% Confidence Intervals) and P heterogeneity (P het) for between group differences. Adjusted geometric means are presented for vitamin B12, D, and triglycerides.
Model 1 was adjusted for age (years) and sex (women, men). Model 2 was model 1 plus BMI (tertials). Model 3 was model 2 plus smoking (never, ever) and alcohol intake (g/day). Vitamin D additionally adjusted for season of measurement (October-March, April-September)
P heterogeneity for significant differences between diet groups obtained via ANCOVA and pairwise testing via Wald's t-tests adjusted for multiple testing using a Bonferroni correction.
a,b,c Pairs of means in the same row with differing letters are significantly different following Bonferroni correction
HDL = High-Density Lipoprotein; LDL = Low-Density Lipoprotein.

**Supplementary Table S4: Sensitivity analysis of adjusted biomarker concentrations across diet groups stratified by supplementation**

| | | | | Omnivorous | Flexitarians | Pescatarians | Vegetarians | Vegans | P for heterogeneity (between group differences) | P for heterogeneity (between supplementation strata) |
|---|---|---|---|---|---|---|---|---|---|---|
| **Vitamin B12 (pmol/L)** | | | **n** | | | | | | | |
| | | Supplement users | **300** | **29** | **40** | **28** | **58** | **145** | | |
| | | Non-Supplement users | **316** | **95** | **88** | **41** | **58** | **34** | | |
| | Model 1 | Supplement users | | 411.0 (342.5, 493.1) | 385.9 (330.0, 451.3) | 401.8 (334.1, 483.2) | 379.6 (331.3, 435.0) | 411.1 (377.2, 448.2) | 0.96 | <0.001 |
| | | Non-Supplement users | | 313.0 (285.6, 343.0) $^{a}$ | 290.1 (262.8, 320.2) $^{a}$ | 301.4 (262.4, 346.1) $^{a}$ | 226.8 (202.0, 254.5) $^{b}$ | 247.6 (213.5, 287.2) $^{a,b}$ | <0.001 | |
| | Model 2 | Supplement users | | 424.0 (354.2, 507.4) | 385.4 (330.5, 449.5) | 408.2 (340.5, 489.5) | 381.9 (334.0, 436.5) | 414.4 (380.8, 451.1) | 0.96 | <0.001 |
| | | Non-Supplement users | | 316.4 (288.5, 346.9) $^{a}$ | 286.8 (259.9, 316.5) $^{a}$ | 299.5 (261.0, 343.7) $^{a}$ | 228.5 (203.6, 256.3) $^{b}$ | 251.3 (216.8, 291.3) $^{a,b}$ | <0.001 | |
| | Model 3 | Supplement users | | 420.9 (351.3, 504.4) | 379.9 (324.9, 444.3) | 405.2 (337.7, 486.2) | 380.8 (332.9, 435.8) | 410.5 (376.4, 447.7) | 0.96 | <0.001 |
| | | Non-Supplement users | | 330.3 (301.1, 362.3) $^{a}$ | 291.6 (264.6, 321.3) $^{a,b}$ | 305.6 (267.3, 349.4) $^{a,b}$ | 239.3 (213.2, 268.6) $^{c}$ | 254.8 (220.8, 294.0) $^{b,c}$ | <0.001 | |
| **Vitamin D (nmol/L)** | | | **n** | | | | | | | |
| | | Supplement users | **414** | **73** | **80** | **46** | **74** | **141** | | |
| | | Non-Supplement users | **176** | **40** | **46** | **21** | **37** | **32** | | |
| | Model 1 | Supplement users | | 72.2 (66.5, 78.4) | 70.8 (65.3, 76.8) | 75.5 (68.1, 83.8) | 72.1 (66.1, 78.5) | 77.8 (73.1, 82.8) | 0.57 | <0.001 |
| | | Non-Supplement users | | 55.5 (48.3, 63.7) | 58.9 (51.4, 67.6) | 48.1 (39.6, 58.4) | 46.5 (40.3, 53.7) | 51.8 (44.4, 60.5) | 0.10 | |
| | Model 2 | Supplement users | | 72.8 (67.0, 79.1) | 70.6 (65.1, 76.5) | 76.4 (68.9, 84.8) | 72.3 (66.3, 78.7) | 77.7 (73.0, 82.7) | 0.56 | <0.001 |
| | | Non-Supplement users | | 54.9 (47.6, 63.4) | 59.7 (51.9, 68.6) | 48.1 (39.5, 58.4) | 46.4 (40.1, 53.6) | 51.3 (43.9, 59.9) | 0.10 | |
| | Model 3 | Supplement users | | 73.1 (67.3, 79.4) | 71.0 (65.4, 77.0) | 76.3 (68.8, 84.7) | 71.9 (65.9, 78.4) | 78.0 (73.2, 83.0) | 0.56 | <0.001 |
| | | Non-Supplement users | | 54.0 (46.4, 62.8) | 59.9 (52.0, 69.0) | 48.5 (39.9, 59.1) | 46.6 (40.2, 54.1) | 51.0 (43.6, 59.7) | 0.10 | |
| **Haemoglobin (g/L)** | | | **n** | | | | | | | |
| | | Supplement users | **84** | **8** | **16** | **4** | **15** | **41** | | |
| | | Non-Supplement users | **146** | **33** | **39** | **19** | **26** | **29** | | |
| | Model 1 | Supplement users | | 135.0 (125.7, 144.3) | 137.5 (131.0, 144.0) | 135.8 (122.6, 149.0) | 135.0 (127.9, 142.1) | 139.7 (135.4, 144.0) | 0.62 | 0.11 |
| | | Non-Supplement users | | 140.0 (136.4, 143.5) | 139.9 (136.4, 143.3) | 137.7 (133.1, 142.3) | 143.3 (139.3, 147.3) | 134.3 (130.4, 138.2) | 0.06 | |

Values are adjusted arithmetic mean values (95% Confidence Intervals) and P heterogeneity (P het) for between group differences. Adjusted geometric means are presented for vitamin B12 and D.
Model 1 was adjusted for age (years) and sex (women, men). Model 2 was model 1 plus BMI (tertials). Model 3 was model 2 plus smoking (never, ever) and alcohol intake (g/day). Vitamin D additionally adjusted for season of measurement (October-March, April-September)
P heterogeneity for significant between diet group differences and between strata of supplementation was obtained via ANCOVA, and pairwise testing via Wald's t-tests adjusted for multiple testing using a Bonferroni correction.
a,b,c pairs of means in the same row with differing letters are significantly different following Bonferroni correction
HDL = High-Density Lipoprotein; LDL = Low-Density Lipoprotein.

**Supplementary Table S5: Sensitivity analysis of adjusted biomarker concentrations across diet groups excluding non-white participants**

| Biomarker | n | Omnivorous | Flexitarians | Pescatarians | Vegetarians | Vegans | P for heterogeneity |
|---|---|---|---|---|---|---|---|
| **Vitamin B12 (pmol/L)** | **591** | **119** | **122** | **68** | **113** | **169** | |
| | | 336.2 (306.7, 368.6) [a,b] | 321.0 (292.5, 352.2) [a,b] | 345.0 (305.7, 389.4) [a,b] | 300.0 (272.5, 330.1) [a] | 375.7 (347.4, 406.3) [b] | 0.008 |
| **Vitamin D (nmol/L)** | **565** | **108** | **120** | **66** | **108** | **163** | |
| | | 65.1 (60.0, 70.8) | 65.2 (60.1, 70.7) | 65.0 (58.4, 72.3) | 61.4 (56.4, 66.8) | 72.0 (67.1, 77.1) | 0.18 |
| **Haemoglobin (g/L)** | **222** | **38** | **52** | **23** | **41** | **68** | |
| | | 140.4 (136.6, 144.1) | 139.2 (135.9, 142.5) | 137.8 (133.1, 142.6) | 140.7 (137.1, 144.4) | 137.2 (134.3, 140.1) | 0.71 |
| **Total Cholesterol (mmol/L)** | **497** | **97** | **104** | **57** | **91** | **148** | |
| | | 5.8 (5.6, 6.0) [a] | 5.6 (5.4, 5.8) [a,b] | 5.4 (5.1, 5.7) [a,b,c] | 5.4 (5.1, 5.6) [b] | 5.0 (4.8, 5.2) [c] | <0.001 |
| **LDL (mmol/L)** | **492** | **96** | **104** | **57** | **89** | **146** | |
| | | 3.3 (3.1, 3.5) [a] | 3.2 (3.0, 3.3) [a,b] | 2.9 (2.7, 3.2) [b,c] | 3.0 (2.8, 3.1) [b] | 2.6 (2.5, 2.8) [c] | <0.001 |
| **HDL (mmol/L)** | **519** | **102** | **107** | **57** | **97** | **156** | |
| | | 1.8 (1.7, 1.9) [a] | 1.8 (1.7, 1.8) [a,b] | 1.8 (1.6, 1.9) [a,b] | 1.6 (1.5, 1.7) [a,b] | 1.6 (1.5, 1.7) [b] | <0.001 |
| **Triglycerides (mmol/L)** | **497** | **97** | **104** | **57** | **91** | **148** | |
| | | 1.4 (1.3, 1.5) [a] | 1.4 (1.3, 1.5) [a] | 1.5 (1.3, 1.6) [a] | 1.6 (1.4, 1.7) [a] | 1.5 (1.4, 1.6) [a] | 0.05 |
| **Non-HDL (mmol/L)** | **497** | **97** | **104** | **57** | **91** | **148** | |
| | | 4.0 (3.8, 4.2) [a] | 3.8 (3.6, 4.0) [a] | 3.6 (3.4, 3.9) [a,b] | 3.7 (3.5, 3.9) [a,b] | 3.4 (3.2, 3.5) [b] | <0.001 |
| **Total Cholesterol: HDL (mmol/L)** | **497** | **97** | **104** | **57** | **91** | **148** | |
| | | 3.6 (3.4, 3.8) [a] | 3.3 (3.1, 3.6) [a] | 3.2 (2.9, 3.5) [a] | 3.5 (3.3, 3.7) [a] | 3.3 (3.1, 3.4) [a] | 0.01 |
| **Triglycerides: HDL (mmol/L)** | **497** | **97** | **104** | **57** | **91** | **148** | |
| | | 1.1 (0.9, 1.4) | 1.0 (0.7, 1.2) | 1.0 (0.7, 1.3) | 1.2 (0.9, 1.4) | 1.2 (1.0, 1.3) | 0.43 |

Values are adjusted arithmetic mean values (95% Confidence Intervals) and P heterogeneity (P het) for between group differences. Adjusted geometric means are presented for vitamin B12 and D.

Model 1 was adjusted for age (years) and sex (women, men). Vitamin D additionally adjusted for season of measurement (October-March, April-September)

P heterogeneity for significant between diet group differences obtained via ANCOVA, and pairwise testing via Wald’s t-tests adjusted for multiple testing using a Bonferroni correction.

a,b,c Pairs of means in the same row with differing letters are significantly different following Bonferroni correction

HDL = High-Density Lipoprotein; LDL = Low-Density Lipoprotein.

**Supplementary Table S6: Sensitivity analysis of adjusted biomarker concentrations across diet groups additionally adjusted for transit time.**

| Biomarker | n | Omnivorous | Flexitarians | Pescatarians | Vegetarians | Vegans | P for heterogeneity |
|---|---|---|---|---|---|---|---|
| **Vitamin B12 (pmol/L)** | **614** | **123** | **128** | **69** | **116** | **178** | |
| | | 336.3 (307.0, 368.5) [a,b] | 323.2 (295.3, 353.7) [a,b] | 346.8 (307.7, 390.9) [a,b] | 301.2 (274.0, 331.0) [a] | 374.5 (347.0, 404.2) [b] | 0.01 |
| **Vitamin D (nmol/L)** | **588** | **112** | **126** | **67** | **111** | **172** | |
| | | 65.2 (60.1, 70.8) | 65.1 (60.2, 70.5) | 65.2 (58.7, 72.5) | 62.1 (57.1, 67.5) | 72.3 (67.6, 77.3) | 0.20 |
| **Haemoglobin (g/L)** | **230** | **41** | **55** | **23** | **41** | **70** | |
| | | 138.4 (134.7, 142.1) | 139.0 (135.8, 142.2) | 137.4 (132.7, 142.1) | 140.5 (136.9, 144.1) | 137.0 (134.2, 139.9) | 0.82 |
| **Total Cholesterol (mmol/L)** | **517** | **99** | **110** | **58** | **93** | **157** | |
| | | 5.8 (5.6, 6.0) [a] | 5.6 (5.4, 5.8) [a,b] | 5.4 (5.1, 5.7) [a,b,c] | 5.4 (5.2, 5.6) [b,c] | 5.0 (4.8, 5.2) [c] | <0.001 |
| **LDL (mmol/L)** | **511** | **98** | **110** | **58** | **91** | **154** | |
| | | 3.3 (3.2, 3.5) [a] | 3.2 (3.0, 3.3) [a,b] | 2.9 (2.7, 3.1) [b,c] | 3.0 (2.8, 3.1) [b] | 2.6 (2.5, 2.8) [c] | <0.001 |
| **HDL (mmol/L)** | **539** | **104** | **113** | **58** | **99** | **165** | |
| | | 1.8 (1.7, 1.9) [a] | 1.8 (1.7, 1.8) [a] | 1.8 (1.7, 1.9) [a] | 1.6 (1.5, 1.7) [a,b] | 1.6 (1.5, 1.7) [b] | <0.001 |
| **Triglycerides (mmol/L)** | **517** | **99** | **110** | **58** | **93** | **157** | |
| | | 1.4 (1.3, 1.5) [a] | 1.4 (1.2, 1.5) [a] | 1.4 (1.3, 1.6) [a] | 1.6 (1.4, 1.8) [a] | 1.5 (1.4, 1.7) [a] | 0.03 |
| **Non-HDL (mmol/L)** | **517** | **99** | **110** | **58** | **93** | **157** | |
| | | 4.1 (3.8, 4.3) [a] | 3.8 (3.6, 4.0) [a] | 3.6 (3.4, 3.9) [a,b] | 3.7 (3.5, 4.0) [a,b] | 3.4 (3.2, 3.6) [b] | <0.001 |
| **Total Cholesterol: HDL (mmol/L)** | **517** | **99** | **110** | **58** | **93** | **157** | |
| | | 3.6 (3.4, 3.8) [a] | 3.3 (3.1, 3.6) [a] | 3.2 (2.9, 3.5) [a] | 3.5 (3.3, 3.7) [a] | 3.3 (3.1, 3.5) a | 0.01 |
| **Triglycerides: HDL (mmol/L)** | **517** | **99** | **110** | **58** | **93** | **157** | |
| | | 1.1 (0.9, 1.4) | 0.9 (0.7, 1.2) | 1.0 (0.6, 1.3) | 1.2 (0.9, 1.4) | 1.2 (1.0, 1.4) | 0.30 |

Values are adjusted arithmetic mean values (95% Confidence Intervals) and P heterogeneity (P het) for between group differences. Adjusted geometric means are presented for vitamin B12 and D.

Model was adjusted for age (years), sex (women, men), and sample transit time (hours). Vitamin D additionally adjusted for season of measurement (October-March, April-September)

P heterogeneity for significant between diet group differences obtained via ANCOVA, and pairwise testing via Wald's t-tests adjusted for multiple testing using a Bonferroni correction.

a,b,c Pairs of means in the same row with differing letters are significantly different following Bonferroni correction

HDL = High-Density Lipoprotein; LDL = Low-Density Lipoprotein.
